# Relative Improvement in Language vs Motor Functions with Reperfusion Therapies for Acute Stroke due to Large Vessel Occlusion

**DOI:** 10.1101/2024.06.27.24309619

**Authors:** Zaka Ahmad, Vivek Yedavalli, Wladimir Sarmiento Gonzalez, Argye E. Hillis

**Affiliations:** Howard University Hospital, Department of Neurology; Johns Hopkins School of Medicine, Department of Radiology; Johns Hopkins University Krieger School of Arts and Sciences; Departments of Neurology and Physical Medicine & Rehabilitation, Johns Hopkins School of Medicine

## Abstract

**Background:** When weighing potential risks versus benefits of reperfusion therapy, the functions likely to recover if blood flow can be restored should be considered. Because deep and motor areas of the brain often infarct relatively early in acute stroke, we hypothesized that reperfusion therapies are more likely to improve language function and neglect (cortical functions) more than motor function.

**Methods:** In this retrospective review of a prospectively collected database, patients with acute stroke due to large vessel occlusion), we evaluated percent improvement (mean change in score/maximum score) for different items of the National Institutes of Health Score Scale with and without endovascular thrombectomy, and/or intravenous thrombolysis.

**Results:** In total, 290 patients (mean age 61.8; SD 14.0; 47.9% female) met the inclusion criteria. For all outcome measures (percent change in language, total language, motor, and neglect) there were significant effects of treatment group (p<0.0001 for all), with the greatest change in the EVT +tPA group, then EVT only group, followed by tPA only, followed by no intervention. Differences between EVT + tPA and EVT only were not significant (p=.30 to 0.79 across outcomes). For patients with aphasia and/or right sided weakness before treatment, the percent change in language was significantly greater than the percent change in weakness (29.8% vs. 12.7%; t(93)=5.3;p<0.0001). Greater percent improvement in language was observed in all treatment groups (p=0.0003 to 0.03 across treatment groups).

**Conclusions:** In patients with acute ischemic stroke due to LVO, improvements in all neurological functions occur with tPA, and even more with EVT (with and without IV tPA). However, gains in language are even greater than gains in motor function with both interventions. Few patients had neglect before treatment, but of those who did, the majority improved, and most (92.8%) improved with EVT.

## Introduction

Endovascular thrombectomy (EVT) and intravenous thrombolysis (tPA) are effective treatments for acute ischemic stroke secondary to large vessel occlusion (LVO). However, they also carry some risks. When clinicians, patients, and their caregivers weigh the potential risks versus benefits of reperfusion therapy, they should consider what functions are likely to recover if blood flow can be restored. Because deep and motor areas of the brain, including caudate, putamen, insular ribbon, middle frontal gyrus, frontal lobe subcortical white matter, precentral gyrus, and frontal lobe paracentral lobule are particularly vulnerable to hypoperfusion and infarct relatively early in acute stroke,^1^ we hypothesized that reperfusion therapies are more likely to improve language function and neglect (cortical functions), more than motor function. That is, successful restoration of blood flow might be more likely to salvage areas that are not already ischemic, including areas critical for language^2–4^ and spatial attention,^5–7^ such as left inferior frontal gyrus, temporal cortex, and inferior parietal cortex. The goal of our study is to evaluate the percent improvement (mean change in score/maximum score) for different items of the National Institutes of Health Score Scale (NIHSS) with and without EVT, and/or tPA in series of patients who had LVO, pretreatment CT angiogram (CTA) to confirm LVO, and NIHSS for evaluation of acute ischemic stroke (a convenience sample from three hospitals).

## Methods

### Participants

In this IRB approved retrospective study of our prospective collected database, patients with AIS caused by an LVO (defined as distal internal artery, M1, or proximal M2 middle cerebral artery segments) on CTA from 2017-2022 from three centers within our larger hospital enterprise. This study was approved through the Johns Hopkins School of Medicine institutional review board (JHU-IRB00269637) and follows the STROBE checklist guidelines as an observational study.

### Statistical Analysis

We first evaluated percent change (change in score/maximum score) in language, total language (language item + orientation questions and commands), motor (strength in arms and legs and face) across treatment groups (tPA only, EVT only, EVT plus tPA, and no reperfusion therapy) using ANOVA. For all further analyses, we combined EVT only and EVT plus tPA, because there were only small differences between these two groups in percent change in the items of interest. We then used paired t-tests to evaluate differences in percent change in total motor function versus the language item (and total language score) for participants who were aphasic (1 or more points on the language item) and/or had right sided weakness of the arm, leg, or face (1 or more points). We also used paired t-tests to evaluate differences in percent change in total motor function versus neglect/extinction for participants who were had neglect/extinction (1 or more points on this item) and/or had weakness of the left arm, leg, or face (1 or more points). Finally, we carried out Fisher’s exact tests to evaluate the association between improvement (dichotomous value as 0 and >0 points change and each treatment. P value of =< 0.05 was considered significant.

### Data Availability

Anonymized data not published within this article will be made available by request from any qualified investigator.

## Results

A total of 290 patients with LVO were included in the analyses. Mean age was 61.8 (SD 14.0; range 18-97); 139 (47.9%) were female. MRI confirmed infarct was in the left hemisphere in 150 (51.7%) and right hemisphere in 140 (48.3%).

Of the 290 patients, 37 (12.8%) received tPA only; 21 (7.2%) underwent EVT and tPA; 18 (6.2%), and 214 (73.8%) received neither treatment. The mean NIHSS score was 4.7 (SD 4.4) in the tPA group, 11.8 (SD 7.1) in the EVT plus tPA group, 10.8 (SD 7.0) in the EVT only group, and 2.7 (SD 4.4) in the group who received neither intervention.

For the entire population, there were significant differences between treatment groups for all outcome measures (Table 1). For all outcome measures (percent change in language, total language, motor, and neglect) there were significant effects of treatment group (p<0.0001 for all), with the greatest change in the EVT+tPA only group, then EVT only group, followed by tPA only, followed by no intervention. For remaining analyses, we combine EVT only and EVT + tPA, since the differences were small and non-significant (by t-test) for all outcomes. Likewise, hereafter, we also report results for language item alone, rather than total language, as there was generally less change in orientation and simple commands, but no significant difference between the percent improved in the two outcomes with tPA (p=0.80) or EVT (p=0.70).

**Table 1:** Percent Change in Each Function and Total Change in NIHSS Score for All Patients.

|  | Mean (*SD*) Change |  |  |  |  |
| --- | --- | --- | --- | --- | --- |
|  | Language Item | Total Language | Neglect | Motor | NIHSS Score |
| **tPA Only** (N=37) | 14.4%<br>(30.0%) | 7.8%<br>(17.9%) | 9.5%<br>(23.1%) | 5.8%<br>(15.3%) | 2.84<br>(0.70) |
| **EVT+tPA** (N=21) | 31.7%<br>(37.2%) | 27.0%<br>(33.1%) | 16.7%<br>(28.9%) | 27.1%<br>(20.1%) | 9.86<br>(1.39) |
| **EVT Only** (N=18) | 22.2%<br>(36.2%) | 20.4%<br>(28.1%) | 19.4%<br>(34.9%) | 20.8%<br>(16.6%) | 9.00<br>(1.54) |
| **No Intervention** (N=214) | 4.7%<br>(18.6%) | 4.1%<br>(15.6%) | 1.9%<br>(10.7%) | 2.5%<br>(9.1%) | 1.40<br>(0.25) |
| **Total** (N=290) | 9.0%<br>(24.6%) | 7.3%<br>(19.7%) | 5.0%<br>(17.7%) | 5.9%<br>(13.8%) | 2.67 |
| **ANOVA:** |  |  |  |  |  |
| *F* | 15.65 | 12.91 | 11.80 | 38.76 | 40.54 |
| *p-value* | <0.0001 | <0.0001 | <0.0001 | <0.0001 | <0.0001 |

For patients with aphasia and/or right sided weakness, the percent change in language was greater than the percent change in weakness (29.8 vs. 12.4; t =5.3; df93; p<0.0001; see Table 2 for 95% confidence intervals). The greater improvement in language than motor function was observed for all treatment groups (Table 2). For those who received tPA the mean difference was 35.4% (SD 35.4) versus 6.6% (SD 14.3) (t=3.1; df15; p=0.008). For those who received EVT (with or without tPA), the difference for percent change in language vs weakness was 46.4% (SD 37.3) versus 28.1% (SD19.0) (t=2.3; df22; p=0.03). For patients who received neither EVT nor IV tPA (the largest group), the difference was 21.2% (SD 29.7) versus 7.5% (SD 15.7) (t=3.9; df54; p=0.0003). Relatively few patients had neglect at baseline, and there was no significant difference in percentage improvement in neglect versus weakness, in those with neglect or weakness pretreatment, with either intervention. However, among the few with neglect and weakness pretreatment, there was greater percent improvement in neglect compared to motor function (66.7 ± 28.9 versus -1.7% ±3.0; t=4.3; df2; p=0.0498).

**Table 2:** Percent Change in each Function Among those with Pre-Treatment Deficits.

|  | **Mean** | **Standard**<br>**Deviation** | **95%**<br>**Confidence**<br>**Interval** |
| --- | --- | --- | --- |
| **Combined Groups (N=94)** |  |  |  |
| Percent Change in Language | 29.8 | 34.0 | 22.8 - 36.8 |
| Percent Change in Motor | 12.4 | 18.5 | 8.6 - 16.1 |
| **tPA Only (N=16)** |  |  |  |
| Percent Change in Language | 35.4 | 35.4 | 16.5 - 54.3 |
| Percent Change in Motor | 6.6 | 14.3 | -1.1 - 14.2 |
| **EVT (with or without tPA) (N=23)** |  |  |  |
| Percent Change in Language | 46.4 | 37.3 | 30.3 - 62.5 |
| Percent Change in Motor | 28.1 | 19.0 | 20 - 36.3 |
| **No Intervention (N=55)** |  |  |  |
| Percent Change in Language | 21.2 | 29.7 | 13.2 - 29.2 |
| Percent Change in Motor | 7.5 | 15.7 | 3.2 - 11.7 |

Only 26 patients had neglect/extinction pre-treatment; 6/8 (75%) who received tPA showed some improvement in neglect, 13/14 (92.9%) who received EVT showed some improvement with treatment, and 7/10 (70%) showed some improvement with neither intervention (ns by Fisher’s exact).

The association between change in each outcome measure (as a dichotomous value) and treatment group was significant by Fisher’s exact both language and strength. Figure 1 shows the percentage of patients who showed any improvement on language, weakness, or neglect for those who had deficits at baseline. Interestingly, of those with aphasia or right sided weakness at baseline, 62.5% improved in language with tPA, 78.3% improved in language with EVT, and fewer than half (42.9%) improved with neither intervention. For all functions, improvements were greatest with EVT, then tPA, then no treatment. The difference between treatment groups was significant by Fishers Exact for language (p=0.01) and motor function (p<0.0001). A greater percentage of patients showed some improvement in strength than language with EVT and without intervention, while a greater percentage of patients showed some improvement in language than strength with tPA (Figure 1). However, there were no significant differences in percentage of patients who made any improvement in language or any improvement in strength, of those who were aphasia and/or weak pre-treatment, for any of the intervention groups (by Fisher’s exact).

**Figure 1.**
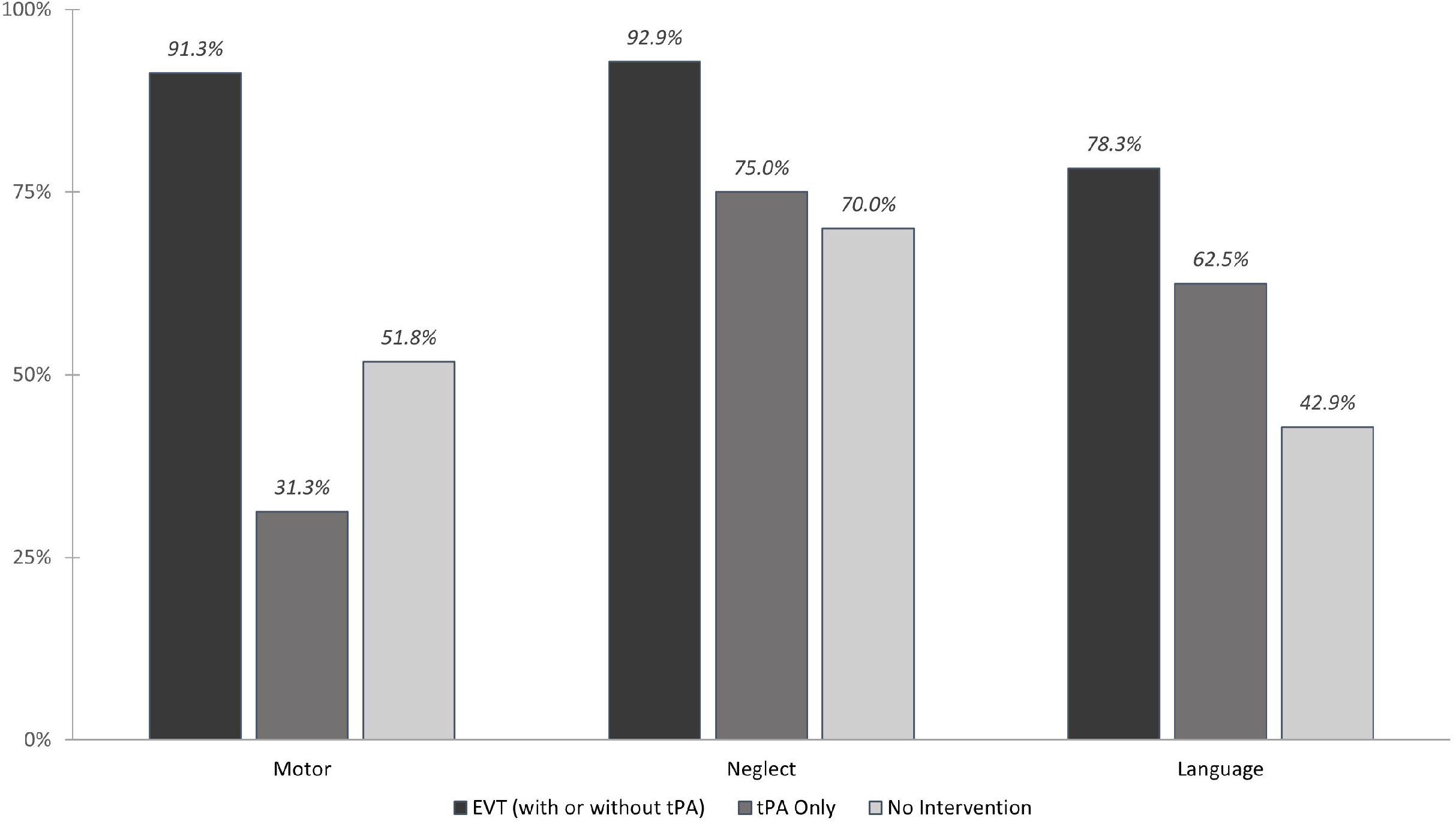
Percentage of Patients Who Improved in Each Function with Each Treatment (of those with Pre-Treatment Deficits)

## Discussion

Consistent with our hypothesis, the cortical function of language improved more (measured as percent change in score) than strength in all treatment groups. Unsurprisingly, EVT had an enormous effect on language, motor, and spatial attention functions; and thrombolysis had a significant, but smaller effect. However, among those with no intervention aimed to restore blood flow, a higher percentage of patients with deficits at bas improved in strength than in language (or spatial attention). Therefore, it is unlikely that results can be accounted for by more a more sensitive assessment item for language than for motor strength. Rather, results may reflect that reperfusion achieved with EVT or tPA (or both), affects cortical functions such as language more than strength. While reperfusion of the motor strip would also improve motor function (as reflected in strong effects of treatment on motor function), early infarct of subcortical tissues may limit the percent improvement in strength.

Our results are important for weighing the risks and benefits of interventions and for counseling regarding prognosis. Patients and families should be advised that deficits such as aphasia might improve more than hemiplegia with intervention, although patients are likely to show at least some improvement in strength with or without treatment (albeit more with treatment).

This information may also have implications for selecting outcome measures for interventions to restore blood flow in acute stroke. Currently, the most common outcome measure is the modified Rankin Scale (mRS). However, the mRS is not especially sensitive to deficits that may improve most, such as aphasia. It is more sensitive to motor functions that impede walking.

There are important limitations to this study. We used items on the NIHSS as the only assessment of language, spatial attention, and strength. While the scoring is reliable, they are very limited measures of all of these functions. It is possible that fine motor control or other motor function might improve more than proximal strength measured by holding up each arm for 10 sec and each leg for 5 sec.

Neglect/extinction was uncommon as measured with the NIHSS, and so we could not compare percent improvement in this domain to other domains. A previous study showed that change in a more objective measurement of neglect (with simple line cancellation) actually correlated better with change in volume hypoperfusion than did change in the total NIHSS score in patients with right hemisphere stroke.^8^ This study was also an observational study, retrospectively analyzing data from a prospectively collected sample of patients. Intervention was not randomized, but was generally determined by following American Heart Association guidelines for treatment of acute ischemic stroke.

Nevertheless, results provide novel and important information about the likelihood and estimated degree of improvement in gross measures of aphasia, neglect, and weakness in acute ischemic stroke, which may be useful in clinical decision-making. In patients with both aphasia and weakness pretreatment, language is likely to improve to a greater degree than weakness with tPA or EVT.

## Acknowledgements

The authors are grateful to the patients who participated in the study.

